# Maintenance therapy with infliximab or vedolizumab in inflammatory bowel disease is not associated with increased SARS-CoV-2 seroprevalence: UK experience in the 2020 pandemic

**DOI:** 10.1101/2020.12.12.20247841

**Authors:** Colleen G C McGregor, Alex Adams, Ross Sadler, Carolina V Arancibia-Cárcamo, Rebecca Palmer, Tim Ambrose, Oliver Brain, Alissa Walsh, Paul Klenerman, Simon Travis, Nicholas M Croft, James O Lindsay, Jack Satsangi

**Author notes:** **Corresponding author contact information:** Address correspondence to: Dr. Colleen McGregor / Prof. Jack Satsangi, Translational Gastroenterology Unit, Experimental Medicine Division, John Radcliffe Hospital (Level 5), Oxford, OX3 9DU, /.

## Abstract

**Background:** There has been great concern amongst clinicians and patients that immunomodulatory treatments for IBD may increase risk of SARS-CoV-2 susceptibility or progression to severe disease.

**Methods:** Sera from 640 patients attending for maintenance infliximab or vedolizumab infusions between April and June 2020 at the John Radcliffe Hospital (Oxford, UK) and Royal London Hospital (London, UK) were tested using the Abbott SARS-CoV-2 IgG assay. Demographic and clinical data were collated from electronic patient records and research databases.

**Results:** Seropositivity rates of 3.0% (12/404), 7.2% (13/180), and 12.5% (7/56) were found in the Oxford and London adult IBD cohorts and London paediatric IBD cohorts respectively. Seroprevalence rates in the Oxford adult IBD cohort were lower than that seen in non-patient facing health-care workers within the same hospital (7.2%). Seroprevalence rates of the London paediatric IBD cohort were comparable to a contemporary healthy cohort collected at the same hospital (54/396, 13.6%).

**Conclusions:** SARS-CoV-2 seropositivity rates are not elevated in patients with IBD receiving maintenance infliximab or vedolizumab infusions. There is no rationale based on these data for elective interruption of maintenance therapy, and we recommend continuation of maintenance therapy. These data do not address the efficacy of vaccination in these patients.

## Introduction

The management of inflammatory bowel disease (IBD) patients treated with biological therapies targeting the immune system has been a key concern throughout the SARS-CoV-2 pandemic. Early guidance from the UK government on self-isolation (“shielding”) to patients who were immunocompromised through disease or therapy, was based on expert opinion rather than evidence(1). Elective withdrawal of immunosuppressive therapy was not advised due to the associated risks of IBD relapse.

Evidence is now emerging to clarify this issue, defining the risk of immunomodulatory treatments on SARS-CoV-2 acquisition and disease course(2). Outcomes in patients with IBD who acquire COVID-19 have been reported by the SECURE-IBD registry(3). These data suggest relative safety of anti-cytokine monotherapy and complement seroprevalence data from Germany which reached provocative conclusions suggesting that patients on anti-cytokine therapy might have reduced susceptibility to SARS-CoV-2(4). In this study, we analyse the SARS-CoV-2 antibody seroprevalence in patients with IBD receiving systemic anti-TNF therapy (infliximab) or gut-selective immunomodulation with vedolizumab in two UK tertiary IBD centres during the first wave of incidence.

## Methods

### Patient recruitment

In total, 640 patients with IBD receiving maintenance infliximab or vedolizumab infusions at the John Radcliffe Hospital (Oxford) and Royal London Hospital, RLH (London) between 20th April - 26th June 2020 were included. Adult (180) and paediatric (56) patients were included from London. Demographic and clinical data were obtained from electronic patient records and IBD databases (Supplementary Table 1). Key differences between the Oxford and London adult cohorts included ethnicity, smoking, comorbidities, disease type, concomitant thiopurines and biologic; in our dataset, London had significantly greater evidence for deprivation than Oxford (Deprivation score 4(3-6.3) *vs*. 8(6-9.3),p<0.001).

**Table 1.** **1a.** Overall SARS-CoV-2 seroprevalence per cohort. **1b.** Seropositivity vs. biologic and IBD diagnoses. **1c.** Univariable relationships between clinical, socioeconomic and demographic factors with SARS-CoV-2 seropositivity. All odds ratios for univariable logistic regression are given with calculated 95% confidence intervals in parentheses. F=fishers test, otherwise logistic regression, all p values uncorrected (Extended analyses Supp. Table 2).

| **1a.** | **Oxford<sup>†</sup>**<br>**n=404** | **London**<br>**n=180** | **London (Paediatric)<sup>§</sup>**<br>**n=56** |
| --- | --- | --- | --- |
| **Overall seroprevalence** | 3.0% (12) | 7.2% (13) *** | 12.5% (7)<sup>‡</sup> |

| **1b.** | **Oxford** | **CD** | **UC** | **IBD-U** | **Total** |
| --- | --- | --- | --- | --- | --- |
|  | **IFX** | 1/105 (1.0%) | 1/66 (1.5%) | 0/3 (0.0%) | 2/176<sup>‡</sup><br>(1.1%) |
|  | **VDZ** | 4/82 (4.9%) | 6/144 (4.2%) | 0/1 (0.0%) | 10/228<sup>^</sup><br>(4.4%) |
|  | **Total** | 5/187 (2.7%) | 7/210 (3.3%) | 0/4 (0.0%) | 12/404<br>(3.0%) |

|  | **London** | **CD** | **UC** | **IBD-U** | **Total** |
| --- | --- | --- | --- | --- | --- |
|  | **IFX** | 6/85 (7.1%) | 2/31 (6.5%) | 0/2 (0.0%) | 8/118<br>(6.8%) |
|  | **VDZ** | 2/21 (9.5%) | 2/40 (5.0%) | 1/1(100%) | 6/62<br>(8.1%) |
|  | **Total** | 8/106 (7.5%) | 4/71 (5.6%) | 1/3(33.3%) | 13/180<br>(7.2%) |

|  | **London (Paediatric)** | **CD** | **UC** | **IBD-U** | **Total** |
| --- | --- | --- | --- | --- | --- |
|  | **IFX** | 3/29 (10.3%) | 3/16 (18.8%) | 0/3<br>(0.0%) | 6/48<br>(12.5%) |
|  | **VDZ** | 0/0 (0.0%) | 1/7 (4.2%) | 0/1<br>(0.0%) | 1/8<br>(4.4%) |
|  | **Total** | 3/29 (10.3%) | 4/23 (17.4%) | 0/4<br>(0.0%) | 7/56<br>(12.5%) |

| **1c.** | **Oxford** |  | **London** |  | **London (Paediatric)** |  |
| --- | --- | --- | --- | --- | --- | --- |
| **Parameter** | **OR (95% CI)** | **p value** | **OR (95% CI)** | **p value** | **OR (95% CI)** | **p value** |
| Age | 0.99 (0.96-1.03) | 0.78 | 1.01 (0.97-1.04) | 0.61 | 0.90 (0.67-1.24) | 0.5 |
| Sex (Male) | 1.80 (0.47-8.32) | 0.39 | 6.68 (0.95-291.9) | 0.06 | 0.52 (0.07-3.46) | 0.45 |
| Weight | 1.02 (0.99-1.05) | 0.19 | 1.00 (0.96-1.03) | 0.98 | 0.99 (0.94-1.04) | 0.66 |
| Deprivation | 0.95 (0.75-1.24) | 0.68 | 1.01 (0.80-1.25) | 0.91 | 0.87 (0.58-1.19) | 0.42 |
| UC diagnosis | 1.60 (0.40-7.58) | 0.55 | 0.66 (0.14-2.50) | 0.57 | 2.08 (0.31-15.77) | 0.43 |
| VDZ | 3.98 (0.83-37.85) | 0.08 | 1.20 (0.30-4.40) | 0.77 | 1.00 (0.02-10.54) | 1 |
| 5-ASA | 3.39 (0.82-12.83) | 0.05 | 0.35 (0.01-2.55) | 0.47 | 0.32 (0.01-2.98) | 0.41 |
| Comorbidity | 0.22 (0.01-1.54) | 0.19 | 4.59 (1.17-17.44) | 0.01 | 0.00 (0.00-8.44) | 1 |

<sup>†</sup>Control data: Seroprevalence in all Oxford HCW 1128/10,034 (11.2%) and in non-patient facing HCW (administrative staff) 88/1218 (7.2%) were higher (p<0.0001 and p<0.0017 respectively) [Chi squared with Yates correction, acknowledging not stratified for confounders]
<sup>§</sup>Control data: Seroprevalence rates of the London paediatric control group were comparable at 54/396 (13.6%)
\*\*\* Oxford vs. London (adult) seroprevalence P ≤ 0.001
<sup>‡</sup> London adult vs. London paediatric seroprevalence - non-significant
<sup>^</sup>including one 'NA' for diagnoses, <sup>‡</sup>including two 'NAs' for diagnoses
IFX: Infliximab, VDZ: Vedolizumab, CD: Crohn's disease, UC: Ulcerative colitis, IBD-U: Inflammatory bowel disease-unclassified, 5-ASA: 5-aminosalicylic acid

### SARS-CoV-2 antibody detection

Sera from all patients were tested using the Abbott SARS-CoV-2 IgG assay and Architect *i*2000SR system, by staff in Oxford. Samples were interpreted as positive according to the manufacturer’s cut off value of ≥1.4(5).

### Statistical analyses

Analysis was performed in R (v3.6). Univariable analysis was performed with Fisher’s exact test for binary variables, and logistic regression for all other variables. Uncorrected P values are given. Odds ratios for univariable logistic regression are given with calculated CIs. Seroprevalence data were compared with available data from healthy healthcare workers (HCW) in Oxford(6) and from a PHE seroprevalence study in unselected paediatric patients at RLH.

## Results

### No increase in overall SARS-CoV-2 seropositivity in patients with IBD on biologics compared to controls

12/404(3.0%) patients tested positive for SARS-CoV-2 antibodies in Oxford. A higher seroprevalence rate was reported from London patients, 13/180 (7.2%) for adults p*≤*0.0001 and 7/56(12.5%) for children (Table 1). Seroprevalence rates in IBD cohorts were lower than their local healthy controls. Seroprevalence in all Oxford Health Care Workers (HCW, 11.2%) and in non-patient facing HCW (7.2%)(6) were higher (p<0.0001 and p<0.0017 respectively). Seroprevalence rates of the London paediatric control group were comparable at 13.6% (54/396, median age 13.0 yrs.(8.1-16.0), male sex 49%).

We also analysed clinical and demographic characteristics of the entire cohort in order to identify any associations of SARS-CoV-2 positive patients. There were no associations with baseline characteristics, including ethnicity, or deprivation status. In Oxford, a trend toward lower seropositivity was observed in patients on infliximab *vs*. vedolizumab (1.1% *vs*. 4.4%); only 2 anti-TNF treated patients were seropositive (Table 1). These trends were not observed in adults or children in London. Concomitant budesonide or 5-ASA use were associated with higher seropositivity rates, although statistical significance was not reached.

## Discussion

We present the results of the first UK data on SARS-CoV-2 seroprevalence in patients with IBD on biologics at the height of the UK pandemic. We report an overall adult seroprevalence of 3.0% and 7.2% in Oxford and London respectively. We also present the first data on a UK paediatric IBD cohort, with a seroprevalence of 12.5%. The differences between centres are consistent with published adult seroprevalence data at the time of our study. The UK Biobank SARS-CoV-2 Serology Study reported a seroprevalence of 6.6% (5.6-7.6%) in the South East (Oxfordshire) and 10.4% (9.6%-11.2%) in London. Coupled with our local control data (7.2%-Oxford, 13.6%-London (Paediatric)), we demonstrate no increase in seropositivity of patients with IBD on biologics in either centre. Furthermore, when compared to both patient-facing and administrative staff in hospital, seroprevalence rates were lower in IBD.

We observed the lowest rate of seropositivity in Oxford patients with IBD on infliximab. Whilst this was not replicated in the more highly exposed London cohort, it is consistent with the observation and hypothesis of Simon and colleagues(4). The SECURE-IBD data reported, in unadjusted analyses, vedolizumab patients had a higher proportion of severe COVID-19 compared with those on anti-TNFs (7.2% *vs*. 2.2%,p=0.007)(2). It has been postulated that anti-cytokine therapies may ameliorate or abrogate the ‘cytokine storm’ associated with severe COVID-19(7).

The seropositivity rates with 5-ASA use in Oxford are also noteworthy in the context of the hypotheses generated by SECURE-IBD data, where 5-ASA use was associated with severe COVID-19 (OR 1.47,95%CI 1.05–2.07)(2). These finding may be artefactual however, reflecting rather the hypothesised potential protective effect of biologics.

Strengths of this study include its deprivation data and two-centre design, with inclusion of both adult and paediatric IBD populations. All samples, including control data, were collected in the same time period and utilised the same assay for analysis. We recognise limitations of our study; including limited power, incomplete exposure history and the presence of confounders with control data comparisons. Whilst the impact of shielding and social distancing practices observed by patients must be considered when interpreting these seroprevalence rates, we consider these findings to be informative, providing evidence to support the continuation of biologics in the COVID-19 era, especially considering the emerging data of deleterious IBD outcomes in those who have discontinued IBD medications during the pandemic(8).

We cannot comment on the durability of response with a single timepoint or whether seropositive patients are protected from re-infection, and therefore we highlight the need for larger, multicentre, prospective, longitudinal studies such as the ICARUS and CLARITY studies, in order to answer the questions of durability and strength of serological response, immunomodulation on SARS-CoV-2 course and risk stratification. These studies also have potential implications in the prioritisation of vaccine delivery in IBD cohorts.

## Data Availability

Please contact the corresponding authors for access to anonymised data

## Acknowledgements

The authors wish to acknowledge the contributions to this work by Stephanie Jones, Jennifer Hollis, Bessie Cipriano, Irish Lee, Kinnari Naik, Polychronis Kemos (QMUL), Ruth Ayling (Barts Health NHS Trust), David Eyre, Philippa Matthews, Oxford Radcliffe Biobank, James Chivenga and the TGU Biobank, Oxford Biomedical Research Centre, IBD Specialist Nurses, TGU Investigators*, Department of Clinical Biochemistry (John Radcliffe Hospital), and Clarissa Oeser (Public Health England). The authors also wish to thank Jean-Frederic Colombel and Serre-Yu Wong for helpful and constructive discussion. * Dr Tim Ambrose, Dr Carolina V Arancibia-Cárcamo, Dr Adam Bailey, Professor Ellie Barnes, Dr Elizabeth Bird-Lieberman, Dr Oliver Brain, Dr Barbara Braden, Dr Jane Collier, Professor James East, Dr Lucy Howarth, Professor Paul Klenerman, Professor Simon Leedham, Dr Rebecca Palmer, Dr Fiona Powrie, Dr Astor Rodrigues, Professor Alison Simmons, Dr Peter Sullivan, Professor Holm Uhlig, Professor Jack Satsangi, Dr Philip Allan, Dr Timothy Ambrose, Dr Jan Bornschein, Dr Jeremy Cobbold, Dr Emma Culver, Dr Michael Pavlides, Dr Alissa Walsh.

## Ethics

Samples from Oxford patients were collected as a project (ref ORB 20/A054) under the ethical approval of the Oxford Radcliffe Biobank, a research tissue bank that has a favourable opinion from the Oxford C South Central REC, with reference 19/SC/0173. Samples from London patients were collected as a project under the ethical approval of the Digestive Disease Bioresource, Barts Health NHS Trust, a research tissue bank that has a favourable opinion from the Bromley REC, reference 15/LO/2127.

## Authors contributions to manuscript

CGCM- patient recruitment, analysis, drafting of manuscript. AA- analysis, drafting of manuscript. RS – sample analysis, manuscript revision. CVA-C, RP, TA, OB, AW, ST, JOL, NMC – patient recruitment, manuscript revision. PK – manuscript revision. JS – initiation of research, recruitment, drafting and revision of manuscript

## Data availability

Please contact the corresponding authors for access to anonymised data.

*Author names in bold designate shared co-first authorship

