## Supplemental Material for "Maintenance therapy with infliximab or vedolizumab in inflammatory bowel disease is not associated with increased SARS-CoV-2 seroprevalence: UK experience in the 2020 pandemic"

### Supplementary material

**Supplementary Table 1:** *Demographic, socioeconomic and clinical characteristics*

| | Oxford n=404 | London (Adult) n=180 | London (Paediatric) n=56 |
|---|---|---|---|
| **Median age (IQR)** | 44.0 (29.0-60.0) | 31.5 (23.0-44.2) **** | 14.8 (12.7-16.3) |
| **Sex (Male) (n, %)** | 214 (53.0) | 119 (66.1) ** | 32 (57.1%) |
| **Median weight (kg) (IQR)** | 74 (64.0-88.0) | 67.5 (59.0-78.0) **** | 53.6 (44.8-66.1) |
| **Median deprivation score[†] (IQR)** | 8 (6.0-9.3) | 4 (3.0-6.3) **** | 4 (2.8-7.0) |
| ***Ethnicity*** | | | |
| White | 349 (86.4) | 74 (41.1) **** | 14 (25.0) |
| Asian | 14 (3.5) | 63 (35.0) **** | 13 (23.2) |
| Black | 3 (0.7) | 11 (6.1) *** | 7 (12.5) |
| Mixed | 3 (0.7) | 0 (0.0) | 2 (3.6) |
| Other | 5 (1.2) | 10 (5.6) ** | 1 (1.8) |
| Unstated | 30 (7.4) | 21 (11.7) | 19 (33.9) |
| ***Smoking status*** | | | |
| Current (n, %) | 41 (11.7) | 8 (4.4) ** | |
| Ex-smoker (n, %) | 114 (32.6) | 12 (6.7) **** | |
| ***Disease*** | | | |
| Crohn's disease (CD) (n, %) | 188 (46.5) | 106 (58.9) ** | 29 (51.8) |
| Ulcerative colitis (UC) (n, %) | 211 (52.5) | 71 (39.4) ** | 23 (41.1) |
| IBD-U (n, %) | 4 (1.0) | 3 (1.7) | 4 (7.1) |
| **Median disease duration, yrs (IQR)** | 10 (5.0-18.0) | 8.5 (4.8-15.2) | 3.3 (2.1-5.9) |
| **Comorbidity (n, %)** | 116 (28.7) | 32 (17.8) ** | 5 (8.9) |
| ***Medication (n, %)*** | | | |
| Infliximab | 176 (43.6) | 118 (65.6) **** | 48 (85.7) |
| Vedolizumab | 228 (56.4) | 62 (34.4) **** | 8 (14.3) |
| Prednisolone | 12 (3.0) | 8 (4.4) | 6 (10.7) |
| Budesonide | 6 (1.5) | 2 (1.1) | 0 (0.0) |
| Corticosteroid | 18 (4.5) | 10 (5.6) | 6 (10.7) |
| 5-ASA | 73 (18.1) | 33 (18.3) | 18 (32.1) |
| Thiopurine | 101 (25.0) | 70 (38.9) *** | 49 (87.5) |
| MTX | 12 (3.0) | 9 (5.0) | 0 (0.0) |
| MMF | 12 (3.0) | 0 (0.0) * | 0 (0.0) |

P values denote comparison of Oxford vs. London (adult): *P ≤ 0.05, **P ≤ 0.01, ***P ≤ 0.001, ****P ≤ 0.0001, non-significant otherwise

[†] Deprivation score: Derived from the English Indices of Multiple Deprivation (IMD), the score classifies relative deprivation per area. A score of 1 = Most deprived, 10 = Least deprived

*IBD-U:* Inflammatory Bowel Disease-Unclassified, *COPD:* Chronic obstructive pulmonary disease, *CVD:* Cerebrovascular disease, *5-ASA:* 5-aminosalicylic acid, *MTX:* Methotrexate, *MMF:* Mycophenolate mofetil

**Supplementary Table 2:** *Extended univariable relationships between clinical, socioeconomic and demographic factors with SARS-CoV-2 seropositivity. All statistics show 95% confidence intervals in parentheses. F=fishers test, otherwise logistic regression, all p values uncorrected*

| | Oxford | | London (Adult) | | London (Paediatric) | |
|---|---|---|---|---|---|---|
| | OR (95% CI) | P value | OR (95% CI) | P value | OR (95% CI) | P value |
| ***Biologic*** | | | | | | |
| **Vedolizumab (VDZ)[F]** | 3.98 (0.83-37.85) | 0.08 | 1.20 (0.30-4.40) | 0.77 | 1.00 (0.02-10.54) | 1 |
| **Infliximab (IFX)[F]** | 0.25 (0.03-1.20) | 0.08 | 0.83 (0.23-3.38) | 0.77 | 1.00 (0.09-52.49) | 1 |
| ***Demographics*** | | | | | | |
| Age | 0.99 (0.96-1.03) | 0.78 | 1.01 (0.97-1.04) | 0.61 | 0.90 (0.67-1.24) | 0.5 |
| **Sex (Male)[F]** | 1.80 (0.47-8.32) | 0.39 | 6.68 (0.95-291.99) | 0.06 | 0.52 (0.07-3.46) | 0.45 |
| Weight | 1.02 (0.99-1.05) | 0.19 | 1.00 (0.96-1.03) | 0.98 | 0.99 (0.94-1.04) | 0.66 |
| Deprivation | 0.95 (0.75-1.24) | 0.68 | 1.01 (0.80-1.25) | 0.91 | 0.87 (0.58-1.19) | 0.42 |
| **Current smoker[F]** | 0.75 (0.02-5.51) | 1 | 1.90 (0.04-16.94) | 0.46 | - | - |
| ***Ethnicity*** | | | | | | |
| White | 0.78 (0.16-7.54) | 0.67 | 0.89 (0.22-3.23) | 1 | 0.47 (0.01-4.46) | 0.67 |
| Asian | 2.64 (0.06-20.92) | 0.35 | 0.81 (0.18-3.07) | 1 | 2.86 (0.36-20.12) | 0.33 |
| Black | 0.00 (0.00-83.87) | 1 | 0.00 (0.00-5.44) | 1 | 1.19 (0.02-13.05) | 1 |
| Mixed | 0.00 (0.00-83.87) | 1 | - | - | 7.48 (0.09-636.53) | 0.24 |
| Other | 0.00 (0.00-38.81) | 1 | 3.57 (0.33-21.30) | 0.16 | 0.00 (0.00-271.84) | 1 |
| ***Clinical*** | | | | | | |
| **Disease UC[F]** | 1.60 (0.40-7.58) | 0.55 | 0.66 (0.14-2.50) | 0.57 | 2.08 (0.31-15.77) | 0.43 |
| Disease duration | 1.00 (0.95-1.05) | 0.88 | 1.00 (0.91-1.06) | 0.97 | 1.03 (0.72-1.39) | 0.86 |
| ***Comorbidities*** | | | | | | |
| **All comorbidity[F]** | 0.22 (0.01-1.54) | 0.19 | 4.59 (1.17-17.44) | 0.01 | 0.00 (0.00-8.44) | 1 |
| **Cancer[F]** | 0.00 (0.00-30.39) | 1 | 9.65 (0.74-94.17) | 0.04 | - | - |
| **Cardiovascular disease[F]** | 0.00 (0.00-5.91) | 1 | 0.00 (0.00-32.66) | 1 | - | - |
| **Chronic kidney disease[F]** | 0.00 (0.00-180.21) | 1 | 0.00 (0.00-497.12) | 1 | - | - |
| **Chronic liver disease [F]** | 1.39 (0.03-10.35) | 0.54 | 4.49 (0.08-61.09) | 0.26 | 0.00 (0.00-39.27) | 1 |
| **COPD / Asthma[F]** | 0.00 (0.00-3.51) | 0.61 | 4.64 (0.71-22.42) | 0.06 | 0.00 (0.00-39.27) | 1 |
| **CVD[F]** | 0.00 (0.00-53.32) | 1 | - | - | - | - |
| **Diabetes mellitus[F]** | 0.00 (0.00-9.35) | 1 | 0.00 (0.00-9.56) | 1 | - | - |
| **Hypertension[F]** | 0.00 (0.00-4.59) | 1 | 2.22 (0.04-20.85) | 0.41 | - | - |
| **Solid organ transplant[F]** | 0.00 (0.00-180.21) | 1 | 0.00 (0.00-497.12) | 1 | - | - |
| **Surgery[F]** | 1.13 (0.19-4.67) | 0.74 | 2.70 (0.71-10.02) | 0.1 | 2.50 (0.04-37.72) | 0.42 |
| ***Concomitant medications*** | | | | | | |
| **Prednisolone[F]** | 0.00 (0.00-13.02) | 1 | 1.90 (0.04-16.94) | 0.46 | 0.00 (0.00-6.54) | 1 |
| **Budesonide[F]** | 6.95 (0.14-70.43) | 0.17 | 0.00 (0.00-70.42) | 1 | - | - |
| **Corticosteroid[F]** | 2.00 (0.04-15.33) | 0.43 | 1.46 (0.03-12.24) | 0.54 | 0.00 (0.00-6.54) | 1 |
| **5-ASA[F]** | 3.39 (0.82-12.83) | 0.05 | 0.35 (0.01-2.55) | 0.47 | 0.32 (0.01-2.98) | 0.41 |
| **Thiopurine[F]** | 0.27 (0.01-1.87) | 0.31 | 0.45 (0.08-1.83) | 0.26 | 0.84 (0.08-44.85) | 1 |
| **MTX[F]** | 0.00 (0.00-13.02) | 1 | 0.00 (0.00-6.96) | 1 | - | - |
| **MMF[F]** | 0.00 (0.00-13.02) | 1 | - | - | - | - |

***COPD:*** **Chronic obstructive pulmonary disease,** ***CVD:*** **Cerebrovascular disease,** ***5-ASA:*** **5-aminosalicylic acid,** ***MTX:*** **Methotrexate,** ***MMF:*** **Mycophenolate mofetil**
